# Integrating Geostatistical Maps And Transmission Models Using Adaptive Multiple Importance Sampling

**DOI:** 10.1101/2020.08.03.20146241

**Authors:** Renata Retkute, Panayiota Touloupou, Maria-Gloria Basanez, T. Deirdre Hollingsworth, Simon E.F. Spencer

**Author notes:** URL: https://www.ntdmodelling.org.

## Abstract

The Adaptive Multiple Importance Sampling algorithm (AMIS) is an iterative technique which recycles samples from all previous iterations in order to improve the efficiency of the proposal distribution. We have formulated a new statistical framework based on AMIS to sample parameters of transmission models based on high-resolution geospatial maps of disease prevalence, incidence, or relative risk. We tested the performance of our algorithm on four case studies: ascariasis in Ethiopia, onchocerciasis in Togo, HIV in Botswana, and malaria in the Democratic Republic of the Congo.

## 1. Introduction

Geostatistical modelling has been applied to map a range of infectious diseases at high spatial resolution and multinational scale; examples comprise: malaria (1), soil-transmitted helminthiasis (2), leishmaniases (3), onchocerciasis (4), dengue (5), human African trypanosomiasis (6), HIV (7), and diphtheria-pertussis-tetanus vaccine coverage (8). These maps are made by averaging over many spatially continuous surfaces, which are constructed using geo-positioned survey data, ecological covariates and spatial correlations (9; 10; 11).

Transmission dynamic models have successfully been utilized to evaluate the population dynamics of infectious diseases and assess the impact of interventions (12). It is a common practice to estimate transmission model parameters based on geographically located data, but geostatistical maps provide a unique opportunity to parameterize transmission models at national or even continental scales. However, it is challenging to explore the whole parameter space efficiently, especially when many possible parameter combinations can produce similar values of model output. Generating a large number of parameter sets and running simulations based on these parameters would require substantial computational resources.

In this work we develop a statistical framework based on the Adaptive Multiple Importance Sampling algorithm (AMIS; (13)) for effective integration of geostatistical maps and transmission models. We investigate the performance of AMIS on four case studies: the soil-transmitted nematode *Ascaris lumbricoides* (causative of ascariasis, a soil-transmitted helminth) in Ethiopia; the filarial parasite *Onchocerca volvulus* (causative of onchocerciasis or river blindness) in Togo; HIV infection in Botswana, and infection by the protozoan *Plasmodium* parasite (causative of malaria) in the Democratic Republic of the Congo (DRC). The results show that the proposed framework can successfully be applied for integrating geostatistical maps and transmission models. The resulting combined output constitutes a geographical projection illustrating how the map will evolve through time as well as how the algorithm can be extended to sample parameters in the presence of multiple sampling times or post-control data.

## 2. Background and methods

Integrating high-resolution geostatistical maps and disease transmission models requires exploring the whole parameter space efficiently, especially when many possible parameter combinations can produce similar values of model output. We propose a statistical framework based on the Adaptive Multiple Importance Sampling algorithm (13) to target important and under-explored areas of the parameter space based on high-resolution nation-wide maps of infection prevalences.

### 2.1. Adaptive Multiple Importance Sampling

Suppose that we need to sample from a complex target distribution *π*. Importance sampling is based on using weighted samples from a proposal distribution Φ, i.e. *θ*_*j*_ ∼ ϕ. The corresponding importance weights are 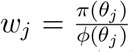, where *π*(*θ*) and *φ*(*θ*) are the target and the proposal density functions (14).

Veach and Guibas (15) proposed the *deterministic multiple mixture* to pool together importance samples from different proposal distributions. In this case, the importance samples 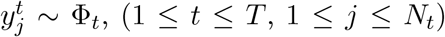 and the corresponding importance weights are calculated based on the mixture of weights (13) given by:

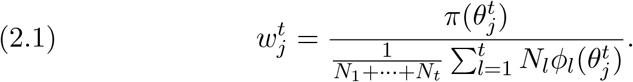

This idea was extended by Cornuet and co-authors (13) who proposed using the deterministic multiple mixture formula to construct importance proposals sequentially and adaptively. Firstly, the importance weights for current iteration, *t*, are calculated, while the importance weights for previous iterations 1 ≤ *u* ≤ *t −* 1 are re-calculated, based on all proposals up to the iteration *t*. Second, samples from all iterations are used to construct the next proposal distribution.

Figure 1 shows differences between importance sampling (IS), adaptive importance sampling (AIS) and adaptive multiple importance sampling (AMIS). In the case of IS, we can reconstruct the target from a proposal distribution by re-weighting sampled parameters according to importance weights, but it may require a lot of samples to have a reasonably good reconstruction. A more efficient scheme is AIS, where samples from previous iterations are used to construct the proposal distribution. As discussed above, AMIS recycles all samples from previous iterations to both construct the proposal for the current iteration, as well as to reconstruct the target distribution.

**Fig 1.**
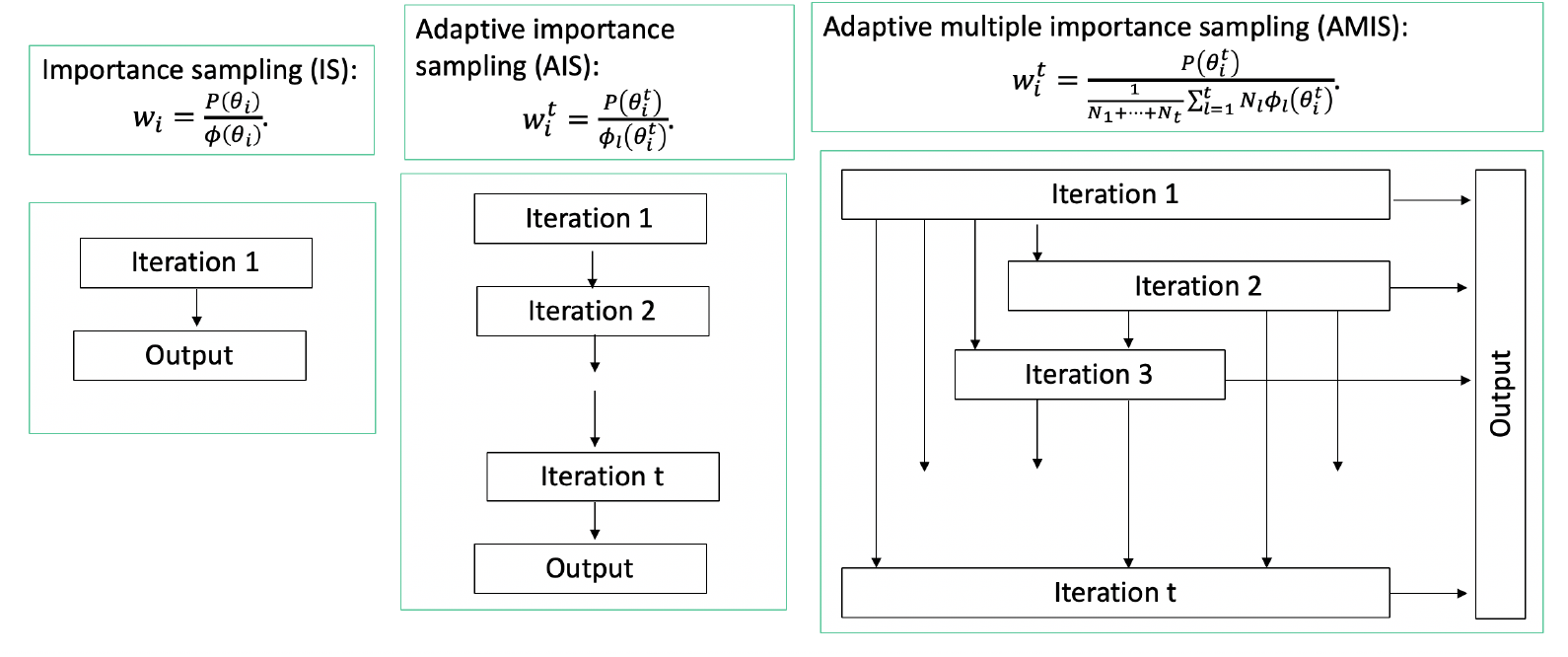
Diagrammatic illustration of differences between importance sampling (left column), adaptive importance sampling (middle column) and adaptive multiple importance sampling (right column).

Adaptive multiple importance sampling has been utilised in a variety of research fields, including population genetics (16), environment illumination computations (17), and signal communications (18).

The novelty of our study is: (i) applying the AMIS approach to real-world geospatial data; (ii) applying AMIS to multiple targets via the same proposal and (iii) working with a ‘moving’ target which changes with each iteration of AMIS. As there is a lack of studies on integrating geostatistical maps and transmission dynamics epidemiological models, we believe that the proposed framework will be a valuable addition to the literature from both theoretical and practical perspective.

### 2.2 Iterative sampling based on a geostatistical map

We assume that a map has *I* pixels (or grid cells), and each pixel has *M* draws of characteristics in which we are interested, e.g. infection prevalences, annual incidence or number of cases. From here onwards, we will assume that we are dealing with the prevalence of infection. Suppose we have an observed prevalence matrix *Q* = (*q*_*i,m*_)_*I*×*M*_, where each row represents a pixel, and each column represents a sampled surface from a geostatistical map.

Projections of infection prevalence can be quantified using a mathematical model (19; 20). Suppose the we have a mathematical model which we define as *F* (*θ*), and this model translates the parameter space onto the prevalence space [0, 1] with individual parameter vector *θ*_*j*_ corresponding to prevalence *p*_*j*_, i.e. *F* (*θ*_*j*_) = *p*_*j*_.

At iteration *t* = 1 parameters are sampled from a prior distribution, then the transmission model is used to calculate the prevalence corresponding to the sampled parameters. Parameter vector *j* is weighted for each pixel *i* so that the weighted distribution of simulated prevalences resembles the distribution of observed prevalences at that pixel. We used Kish’s effective sample size (ESS) (22) as a measure of the quality of representation of the pixel’s prevalence distribution by the simulations.

#### Algorithm AMIS for integrating geostatistical maps and transmission models

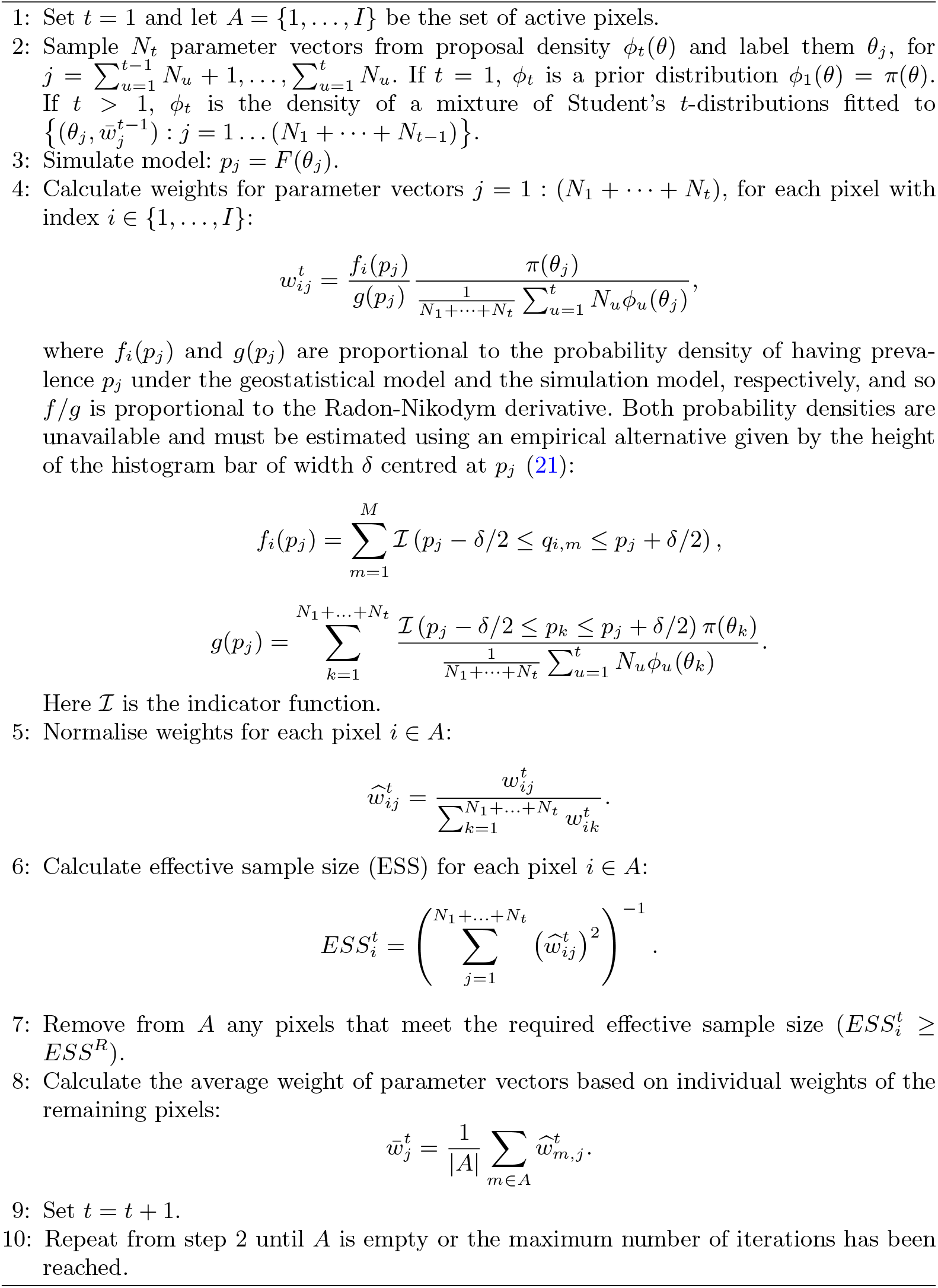

At iterations 2+ we remove pixels which have an ESS above the required threshold *ESS*^*R*^ and then construct a new proposal composed of a mixture of *p*-dimensional Student’s *t*-distributions based on the average weights of the simulations, averaged only over pixels that are yet to reach *ESS*^*R*^. The intuition is that later proposals will be more suitable for pixels that were not well served by earlier proposals. The *t*-distribution has heavier tails than the Gaussian distribution giving a more robust importance proposal and providing capacity to capture a wider range of targets (23). We used the R package mclust to fit the proposal (24). Iterations continue until all pixels have *ESS*_*i*_ *≥ ESS*^*R*^ or the maximum number of iterations reached. Pseudo code is shown in Algorithm AMIS.

### 2.3 Incorporating data from multiple time points

Our proposed framework can be naturally extended to the case where geospatial maps are available at multiple time points *k* = 1 … *K*. In this case, weights for pixel *i* = 1 … *I* pixel, and parameter vector *j* = 1 : (*N*_1_ + … + *N*_*t*_) will be equal to:

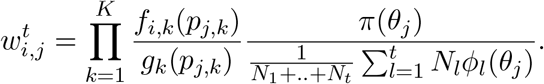

Again, these weights will favour parameter vectors producing simulated projections which are closer to observed prevalences at multiple time points. This approach can be applied to multiple baseline prevalence maps as well as to the incorporation of post-control maps.

## 3. Applications

We applied AMIS to four case studies: ascariasis in Ethiopia, onchocerciasis in Togo, HIV in Botswana, and malaria in the DRC. The framework of AMIS approach and description/objectives of each case study are shown in Figure 2.To set up a cse study, three components are required in order to run AMIS framework: geostatistical map, parameter prior distribution and transmission model. We used survey data (25) to produce a series of prevalence maps of *Ascaris lumbricoides* infection with different spatial resolutions and number of observed prevalences at each pixel. For onchocerciasis, HIV, and malaria, we used publicly available maps (4; 7; 1). We have sampled parameters using the AMIS framework and simulated projections for interventions: mass drug administration (MDA) with ivermectin for onchocerciasis in Togo, and the use of insecticide treated nets (ITNs) for malaria in the DRC. For the latter, we focused only on ITNs because the coverage of artemisinin combination therapies (ACTs) was very low in the DRC (1; 26).

**Fig 2.**
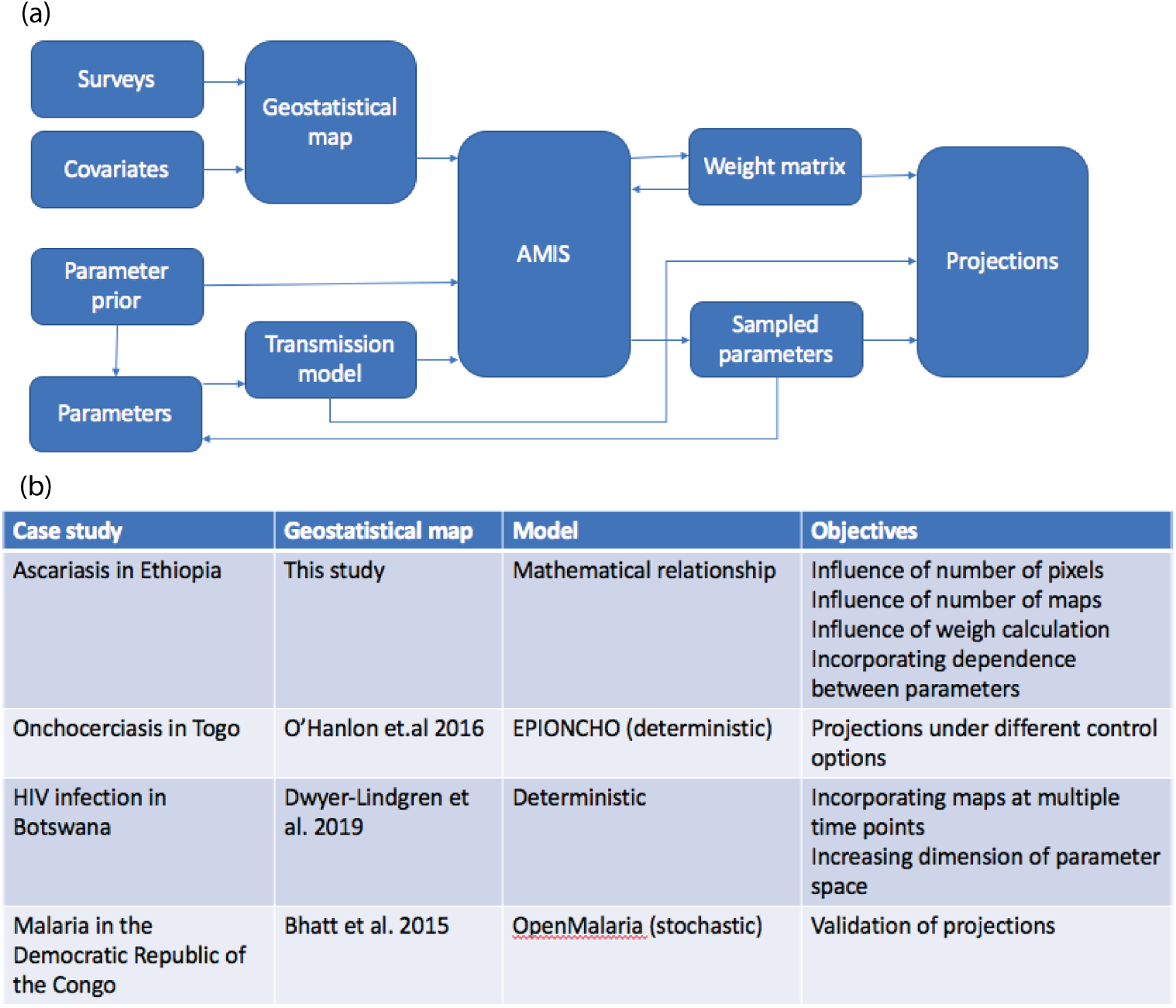
(a) AMIS framework. (b) Description of case studies.

For all four case studies we set *δ* = 0.05, *N*_*t*_ = 1000, *T* = 25, and *ESS*_*R*_ = 2000. Therefore the maximum number of sampled parameter vectors was 25,000.

### 3.1. Ascariasis in Ethiopia

*Ascaris lumbricoides* is an intestinal nematode, also known as roundworm (27). It is estimated that 819 million people worldwide are infected by *A.lumbricoides* (25).

#### 3.1.1. Model and data

In this study we assumed that the mean number of parasites is a parameter, and we use a simple mathematical relationship between the prevalence and the mean intensity of infection, based on fitting a negative binomial distribution to observed data on worm burdens (19),

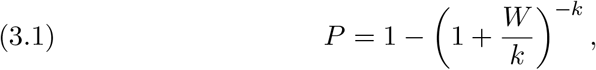

where *W* is the mean number of worms per host, and *k* describes the degree of clumping of parasites within a population of hosts (meaning that the distribution of worms per host is overdispersed compared to a Poisson distribution, with *k* being an inverse measure of overdispersion: the smaller its value, the greater the aggregation of parasites among hosts (28)).

Data on *Ascaris* surveys for Ethiopia were downloaded from the Global Atlas of Helminth Infection (London Applied & Spatial Epidemiology Research Group, LASER) (25). The data consisted of longitude, latitude, number of school-aged children (SAC; 5-14 years old) examined, number of positives (for adult worm presence (25)) and prevalence prior to wide-spread deworming programes (25). Entries with missing geo-location or prevalence values were excluded from further analysis, leaving 290 survey values. We used the INLA-R package (29) and an SPDE approach (30) to produce national level prevalence maps. We incorporated spatial correlation (non-stationary locally isotropic SPDE/GMRF model) and included elevation as an environmental covariate (31) (downloaded from the **raster** package (32)).

#### 3.1.2. Implementation of AMIS

We set a uniform prior for log of the mean number of worms log(*W*) ∼ *U* [log(0.01), log(60)] and a uniform prior for the degree of clumping *k* ∼ *U* [0, 3]. Although typical values for mean worm burden are 10-20 (33), much higher numbers have been observed in field conditions (19; 34).

We produced maps of *Ascaris* prevalence in Ethiopia at resolutions 5km×5km (*I* = 37, 695 pixels) and 10km×10km (*I* = 11, 369 pixels) and sampled 100, 500, 1000 and 2000 individual maps from the posterior distribution for both resolutions. Map showing communities sampled in Ethiopia (25) and triangulated mesh used to build a geostatistical model of *Ascaris* prevalence in Ethiopia (29) are shown in Figure S1.

#### 3.1.3. Results

We compared our proposed framework against an algorithm which samples parameter vectors at each iteration from the prior only, and against a modified AMIS where we set that |*A*| = *M* for all iterations, i.e. algorithm without removing pixels which have achieved *ESS ≥ ESS*^*R*^.

The numbers of samples required to meet the ESS target for all pixels are given in Table 1. It can be seen that the adaptive version of AMIS (i.e. with set of active pixels updated every iteration) outperformed other two methods and required around a third of sampled parameters. Eight iterations were required to achieve min(*ESS*) *≥ ESS*^*R*^ for resolution 10km×10km and *M ≥* 500, corresponding to 8,000 parameter vectors. For the higher resolution map with 5km×5km pixels, the number of parameter vectors increased to 9,000 when the number of maps decreased from *M* = 2, 000 to *M* = 1, 000 and lower. The same number of parameter vectors was required for 10km×10km and *M* = 100.

**Table 1.** Number of iterations to achieve min(ESS) ≥ ESS^R^, with ESS^R^ = 2, 000. Here P stands for sampling from the prior only, A_1_ stands for AMIS with |A| = M for all iterations, and A_2_ is based on algorithm 1. Each iteration samples 1000 parameter sets.

| | $M = 100$ | | | $M = 500$ | | | $M = 1,000$ | | | $M = 2,000$ | | |
| --- | --- | --- | --- | --- | --- | --- | --- | --- | --- | --- | --- | --- |
| $I$ | $P$ | $A_1$ | $A_2$ | $P$ | $A_1$ | $A_2$ | $P$ | $A_1$ | $A_2$ | $P$ | $A_1$ | $A_2$ |
| 11,369 | 23 | 24 | 9 | 21 | 21 | 8 | 22 | 21 | 8 | 21 | 21 | 8 |
| 37,695 | 24 | 18 | 9 | 22 | 18 | 9 | 22 | 18 | 8 | 22 | 17 | 8 |

Figure 3 shows how the number of active pixels |*A*|, minimum and maximum ESS (over pixels) changed with each AMIS iteration for the eight configurations described previously. The AMIS framework produced a consistent efficiency when decreasing the number of active pixels and increasing the minimum ESS, with the exception of *M* = 100 maps. For this configuration both |*A*| and min(*ESS*) had a slight lag in comparison to *M* = 500, *M* = 1000 and *M* = 2000. Although we remove pixels with *ESS ≥ ESS*^*R*^ from the active pixel sets, they are still benefiting from the additionally sampled parameters as can be seen from the max(*ESS*) dynamics.

**Fig 3.**
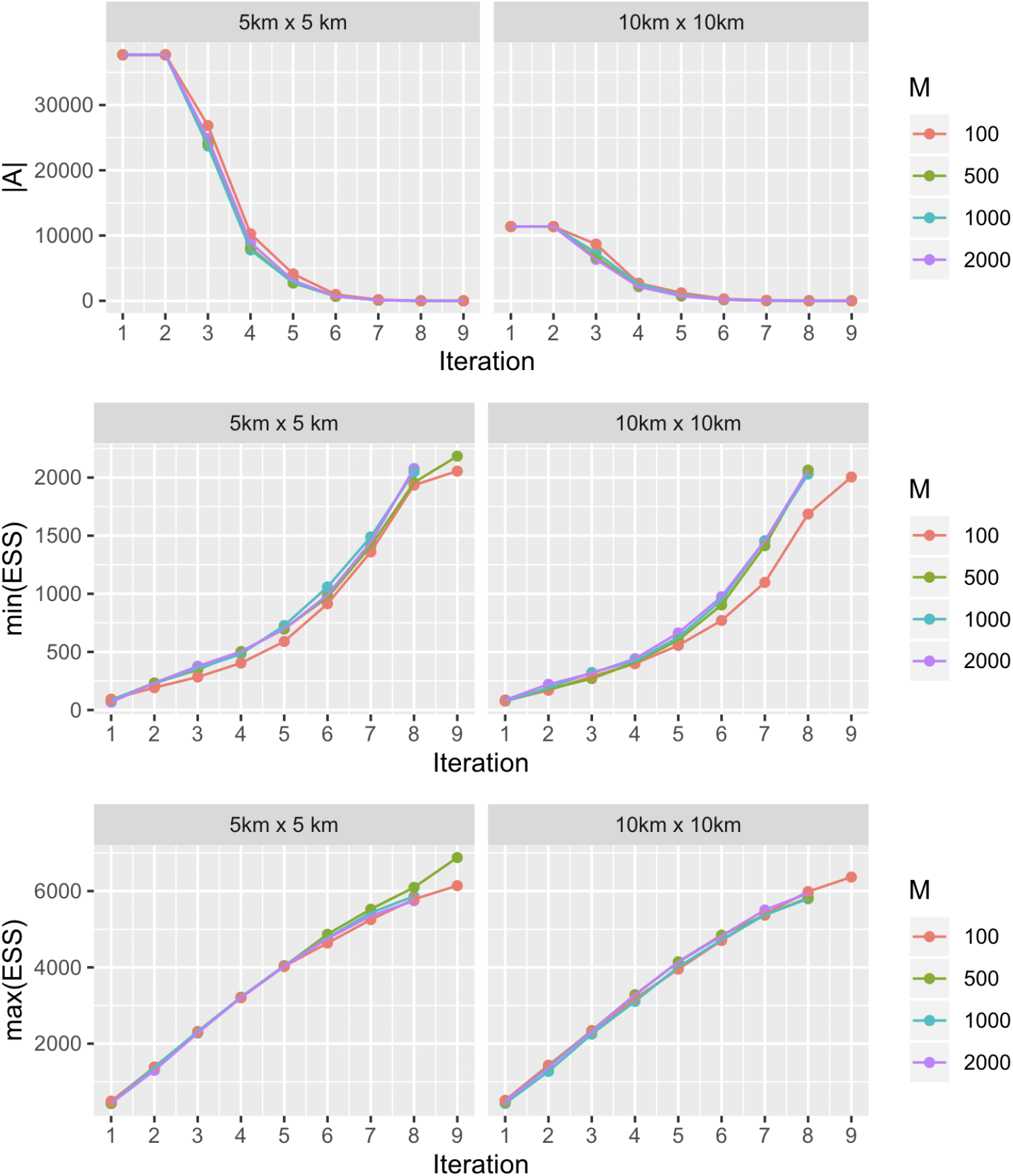
Change in a number of active pixels, minimum of ESS and maximum of ESS with each AMIS iteration plotted as a function of spatial resolution and number of maps for each pixel in the Ascaris lumbricoides case study.

Details of the map and the parameters sampled for resolution 5km×5km and *M* = 2000 are shown in Figure 4. Firstly, we plotted a map of mean prevalence for each pixel (fig.4 (a)), which ranged between 0.1 and 0.55. Second, a histogram of sampled prevalences, colour coded according to a proportion sampled at each iteration, is shown in Fig.4(b). Here, iterations 2 and above targeted prevalences mostly between 0 and 0.5, as this is the range of mapped prevalences of *Ascaris* (Supplementary Figure S1). Third, Fig.4(c) shows prevalence as a function of log mean number of worms, log(*W*), and degree of parasite aggregation, *k*; this function has a complex “L-shaped” dependence with prevalences ranging from 0 to 1 as log(*W*) or *k* increase. Finally, when we look at the density of sampled parameters, the algorithm targeted areas which show the largest variability in terms of simulated prevalences, i.e along this “L-shaped” contour (fig.4 (d)).

**Fig 4.**
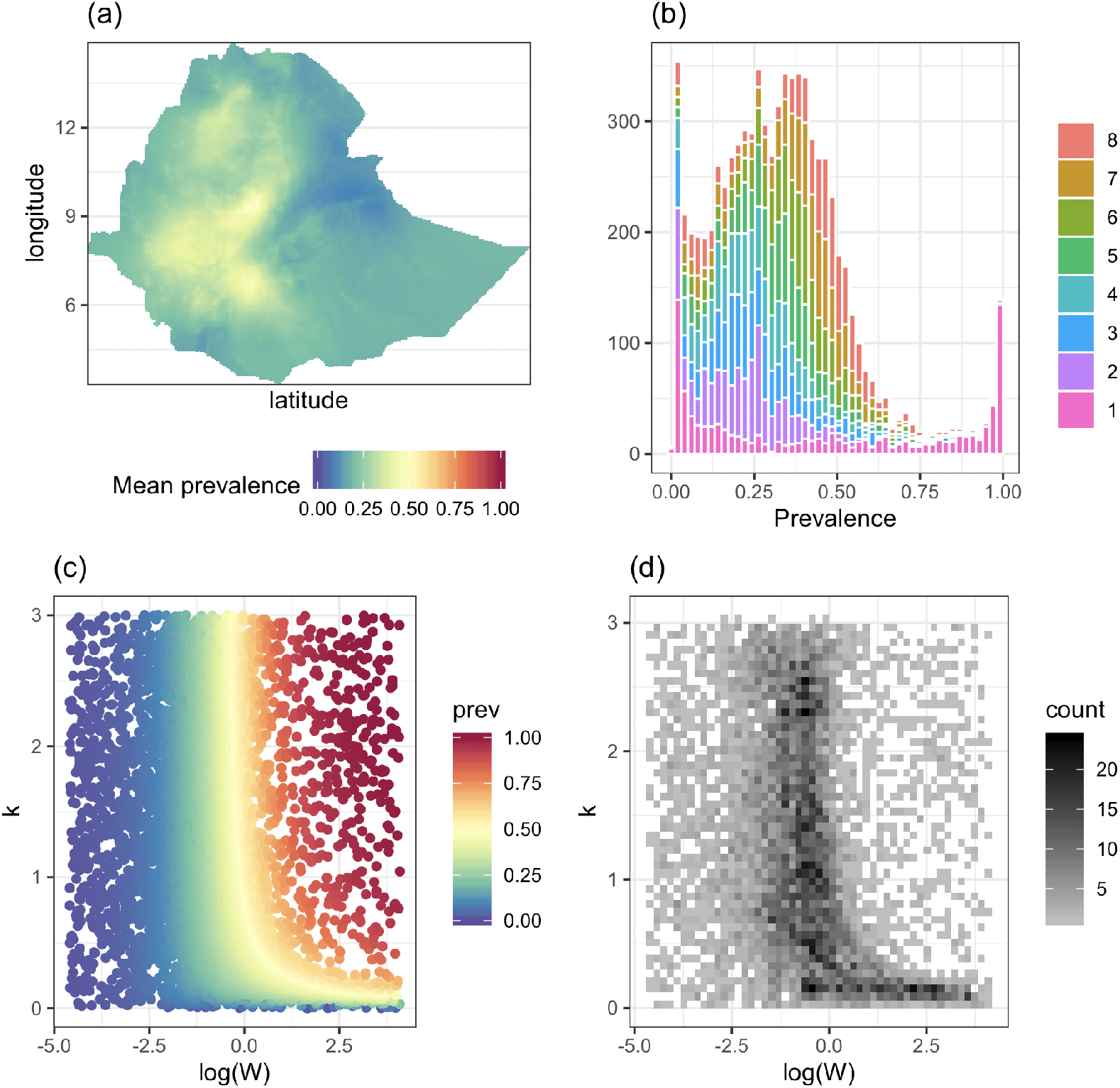
Results for Ascaris lumbricoides in Ethiopia. (a) Map of mean prevalences constructed from survey data in Ethiopia (25). The resolution is 5km×5km and M = 2000. Sampled communities and prevalences are shown in Figure S1. (b) Histogram of sampled prevalences with colour coding according to the iteration of the AMIS algorithm. (c) Scatter plot of sampled model parameters and corresponding prevalence. (d) Density of sampled parameters. Prevalence and density distributions are shown as a function of log mean number of worms, log(W), and degree parasite aggregation, k.

The alternative methods we tested, i.e. based on sampling from the prior only, and based on AMIS with |*A*| = *M* for all iterations, produced samples from different regions of parameter space (Figure 5). When sampling uniformly, we found that a histogram of posterior prevalences has a “U-shape”, i.e. very low and very high values of prevalences are over-sampled, which does not correspond to the mapped prevalences (Figure SI). When sampling with the set of active pixels equal to all pixels, the algorithm targeted parameter regions similar to those of the adaptive AMIS, but still required twice as many iterations.

**Fig 5.**
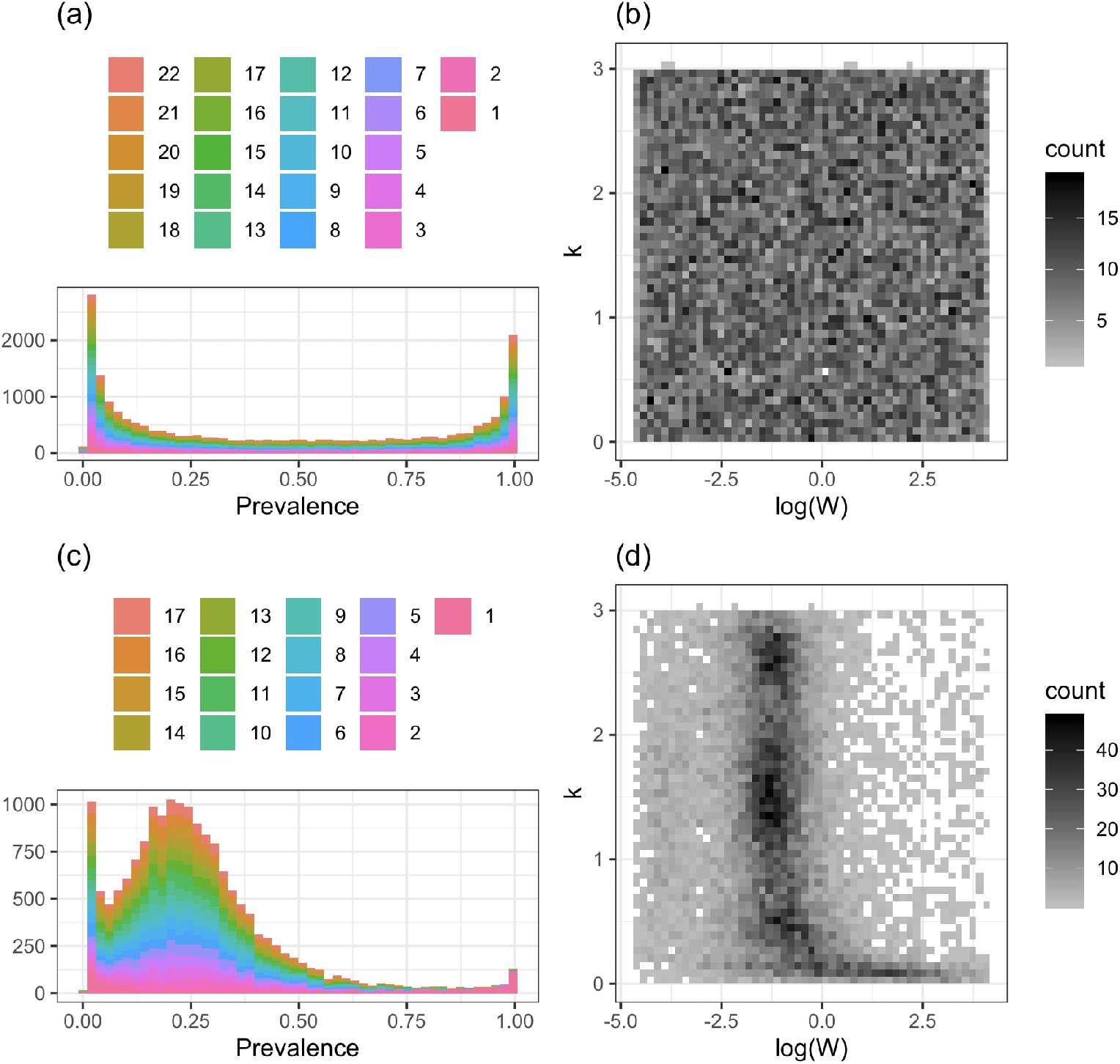
Results for Ascaris lumbricoides in Ethiopia. At each AMIS iteration, the parameters were sampling from the prior (a)-(b); or using weights of all pixels (c)-(d). (a) and shows histogram of sampled prevalences with colour coding according to the iteration of the AMIS algorithm. (c) and (d) shows scatter plot of sampled model parameters and corresponding prevalence. (d) Density of sampled parameters. Prevalence and density distributions are shown as a function of log mean number of worms, log(W), and degree parasite aggregation, k. The resolution is 5km×5km and M = 2000.

#### 3.1.4. Incorporating parameter dependencies

A consistent relationship between mean worm burden and prevalence of *Ascaris lumbricoides* infection has been observed in a data coming from a range of geographically distinct human communities (34). Therefore, we have testing the performance of the AMIS framework when the prior of the parameters incorporates the dependence between mean worm burden, *W*, and degree of parasite aggregation, *k*. We have estimated the relationship between *k* and *W* using paired prevalence and mean intensity data from (34). This have led to the following prior for parameter: uniform prior for log of the mean number of worms log(*W*) ∼ *U* [log(0.01), log(60)] and a Gaussian distribution for the degree of clumping *k* ∼ *N* (0.3337 + 0.0171 ∗ *W, σ*(*W*)) (Supplementary Figure S2). For details on fitting procedure see Appendix A in Supplement.

Figure 6 shows that nine iterations were required for the AMIS algorithm, which is comparable to eight iterations when applying AMIS without dependence between *W* and *k* (Fig. 4).

**Fig 6.**
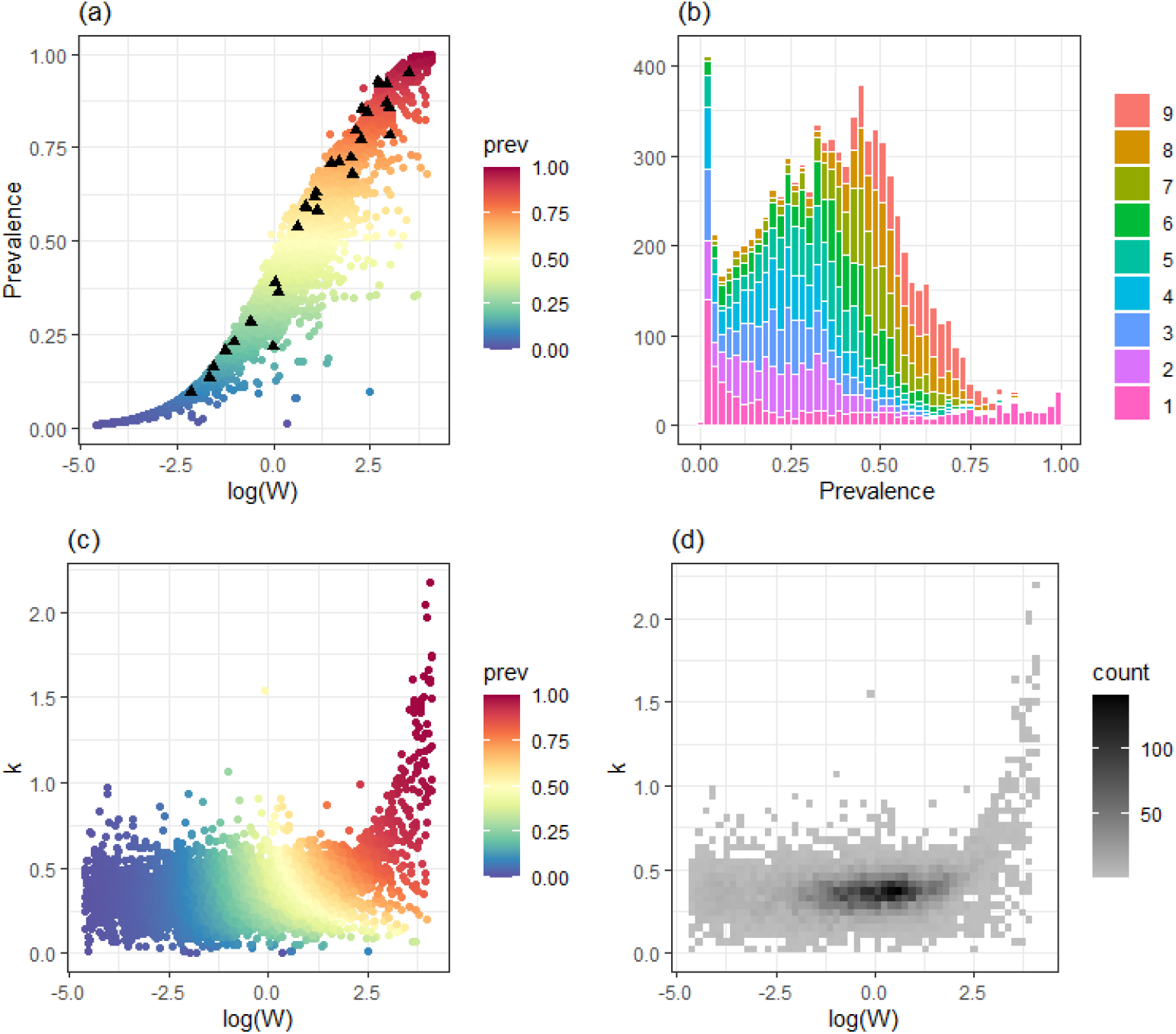
Results for Ascaris lumbricoides in Ethiopia. (a) Sampled values of log mean number of worms, log(W) and corresponding values of prevalence (coloured dots) with the experimental data from (34) (black triangles). (b) Histogram of sampled prevalences with colour coding according to the iteration of the AMIS algorithm. (c) Scatter plot of sampled model parameters and corresponding prevalence. (d) Density of sampled parameters. Prevalence and density distributions are shown as a function of log mean number of worms, log(W), and degree parasite aggregation, k. The resolution is 5km×5km and M = 2000.

### 3.2. Onchocerciasis in Togo

Onchocerciasis is a filarial infection caused by *Onchocerca volvulus* and transmitted among humans via the bites of female *Simulium* blackflies. Onchocerciasis is responsible for skin disease, visual impairment including blindness, and excess mortality, which may be associated with epilepsy (35). Programmes for the control and elimination of the onchocerciasis have been targeted to the areas most affected and great strides have been made, but challenges remain to achieve elimination of transmission (35). Some of these include the lack of efficacious drugs to kill the adult worms (which can live, on average, 10 years, but may live up to 15-20 years (36). Annual or bi-annual mass drug administration (MDA) with ivermectin is the main intervention strategy, but vector control (treating the vector breeding sites in fast flowing rivers) has also been used with success (37). Targeting vector control activities effectively would necessitate an in-depth knowledge of vector breeding site ecology, as well as a better understanding of hydrological conditions in rivers (38; 39; 40).

#### 3.2.1. Model and data

We simulated onchocerciasis transmission in communities using the deterministic version of the EPIONCHO model (41). This model includes: age and sex structure of the human population; age- and sex-specific exposure to blackfly bites; dynamics of the mean number of fertile and non-fertile female worms per host; mean number of microfilariae per mg of skin in the human host and mean number of infective larvae per blackfly vector. All parameters, except the annual biting rate (the number of bites per person per year), *abr*, and *k*, the aggregation of adult warms, were set as in the supplementary material of (41). The annual biting rate determines the intensity of transmission and therefore the prevalence and intensity of infection.

We used maps of baseline (pre-control) microfilarial prevalence (by skin snip microscopy) for ages 5+ years at 5km×5km spatial resolution developed in (4) for the area of the Onchocerciasis Control Programme in West Africa. The data consist of *M* = 2, 000 samples for each pixel, with each individual prevalence map consisting of *I* = 9, 360 pixels.

#### 3.2.2. Implementation of AMIS

We set a uniform prior for the log of the annual biting rate: log(*abr*) ∼ *U* [log(100), log(30, 000)] and a uniform prior for the aggregation of adult warms *k* ∼ *U* [0, 3].

Regular distribution of ivermectin treatment in the form of MDA campaigns is the main current strategy for onchocerciasis control. (Although vector control was also implemented in Togo, we have focused on ivermectin MDA). As ivermectin is mostly microfilaricidal, and the microfilariae are responsible for morbidity and transmission to vectors, this type of intervention reduces disease progression in treated individuals and reduces transmission in the population. For each sampled parameter vector, we have run projections for 15 years under two levels of therapeutic coverage using the EPIONCHO model (41). In particular, we have simulated a coverage of 65% of the total population (the minimum coverage recommended by the World Health Organization), and an enhanced coverage of 80% (as an alternative treatment strategy (42), the latter meaning that treatment is reaching nearly the entirety of the eligible population. In both cases, the proportion of systematic non-adherers was set at 5%.

#### 3.2.3. Results

Similar to the case of ascarisis (Section 3.1), there is a strong heterogeneity in the spatial distribution of onchocerciasis prevalence in Togo (figure 7 (a)). The mean microfilarial prevalence can reach levels of up to 80% in holoendemic areas with high vector biting rates. Figures 7 (b)-(d) show sampled parameter vectors and simulated prevalences. The AMIS framework required 10 iterations to achieve *ESS ≥ ESS*^*R*^ in all pixels. Again, we can see that AMIS sampled most of the parameters along a contour resembling an “L-shape” with quite narrow regions corresponding to the largest variation of prevalences. A histogram of simulated prevalences has a high frequency for values close to zero and two peaks at around 0.25 and 0.8 (Fig.7 (b)).

**Fig 7.**
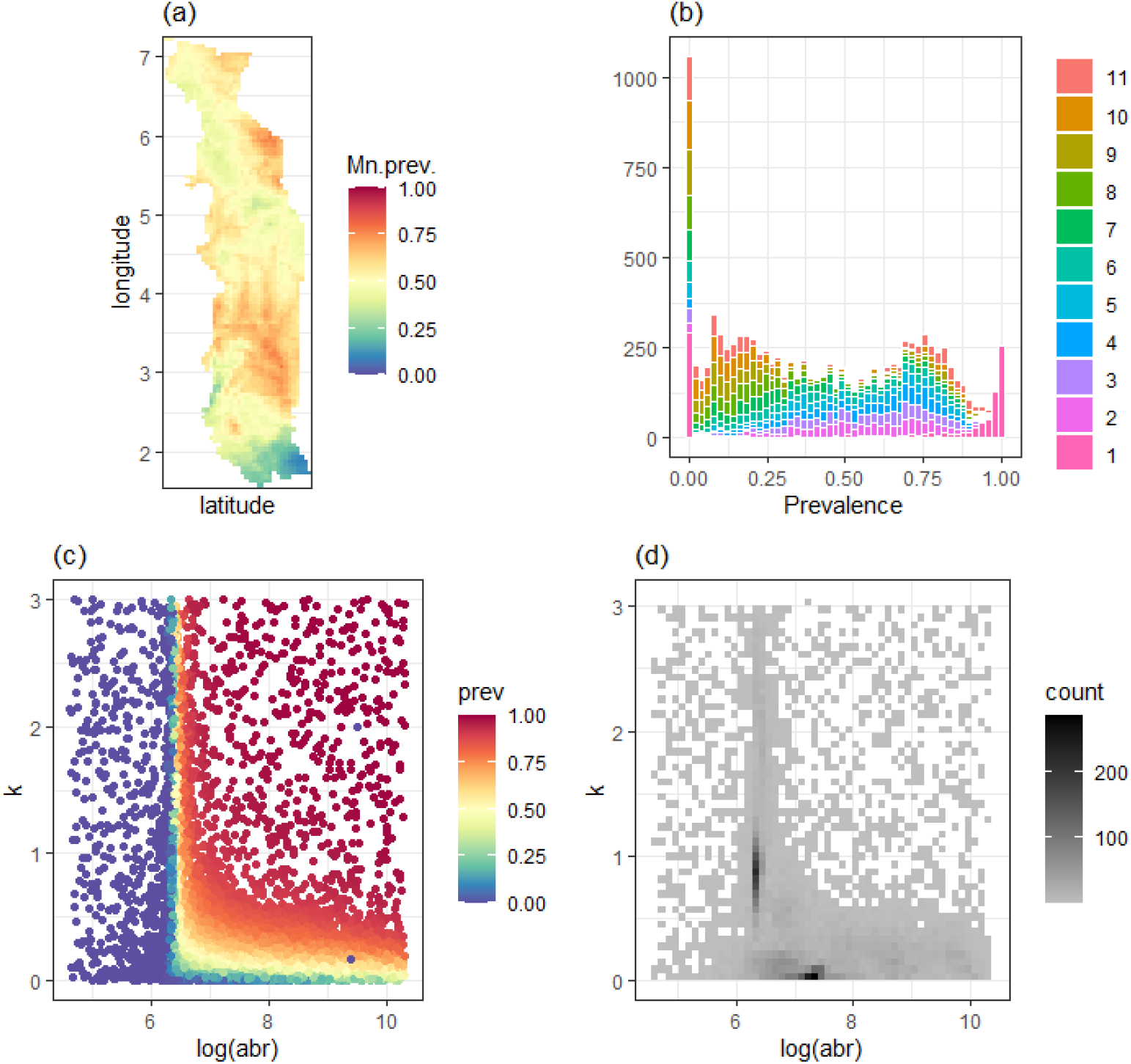
Results for onchocerciasis in Togo. (a) Map of mean baseline prevalence (4). (b) Histogram of sampled prevalences with colour coding according to the various iterations of the AMIS algorithm. (c) Scatter plot of sampled model parameters and corresponding prevalence. (d) Density of sampled parameters. In (c) and (d), prevalence and density distributions are shown as a function of log annual biting rate, log(abr), and the aggregation of adult warms, k.

We compared the impact of 15 years of annual ivermectin MDA with total population coverage 65% and 80%. Figure 8 shows reduction in microfilarial prevalence in endemic area 5, 10 and 15 years after start of MDA. Pre-control endemicity levels (i.e. pre-intervention infection prevalence and intensity) have been indicated as a crucial factor determining the success of intervention strategies to achieve elimination of transmission (42), with areas of higher baseline endemicity indicative of intense transmission (higher basic reproduction ratio) due to high blackfly vector density, where elimination is more difficult (in the absence of vector control). Furthermore, a modelling study using the stochastic version of the model, EPIONCHO-IBM, found that the resilience of the parasite population to MDA was markedly higher for lower levels of exposure heterogeneity *k* (43). Figure 9 shows the temporal dynamics of microfilarial prevalence under annual ivermectin MDA for three pixels corresponding to different levels of mean baseline microfilarial prevalence (0.8 for a hyperendemic/holoendemic situation; 0.5 for a mesoendemic setting, and 0.3 for a hypoendemic scenario). First, it can be seen that there is a good agreement between the mapped and simulated baseline prevalences. Second, uncertainty is propagated with time and simulations produce a wide range of projected prevalences after 15 annual rounds of MDA, especially for the hyperendemic scenario. This is due to equal sampling from different regions of the parameter space, where sets of *{abr, k}* match similar levels of baseline prevalence but have their own characteristic resilience to MDA. Uncertainty can be reduced by utilizing post-control maps within the AMIS algorithm (see Section 3.4).

**Fig 8.**
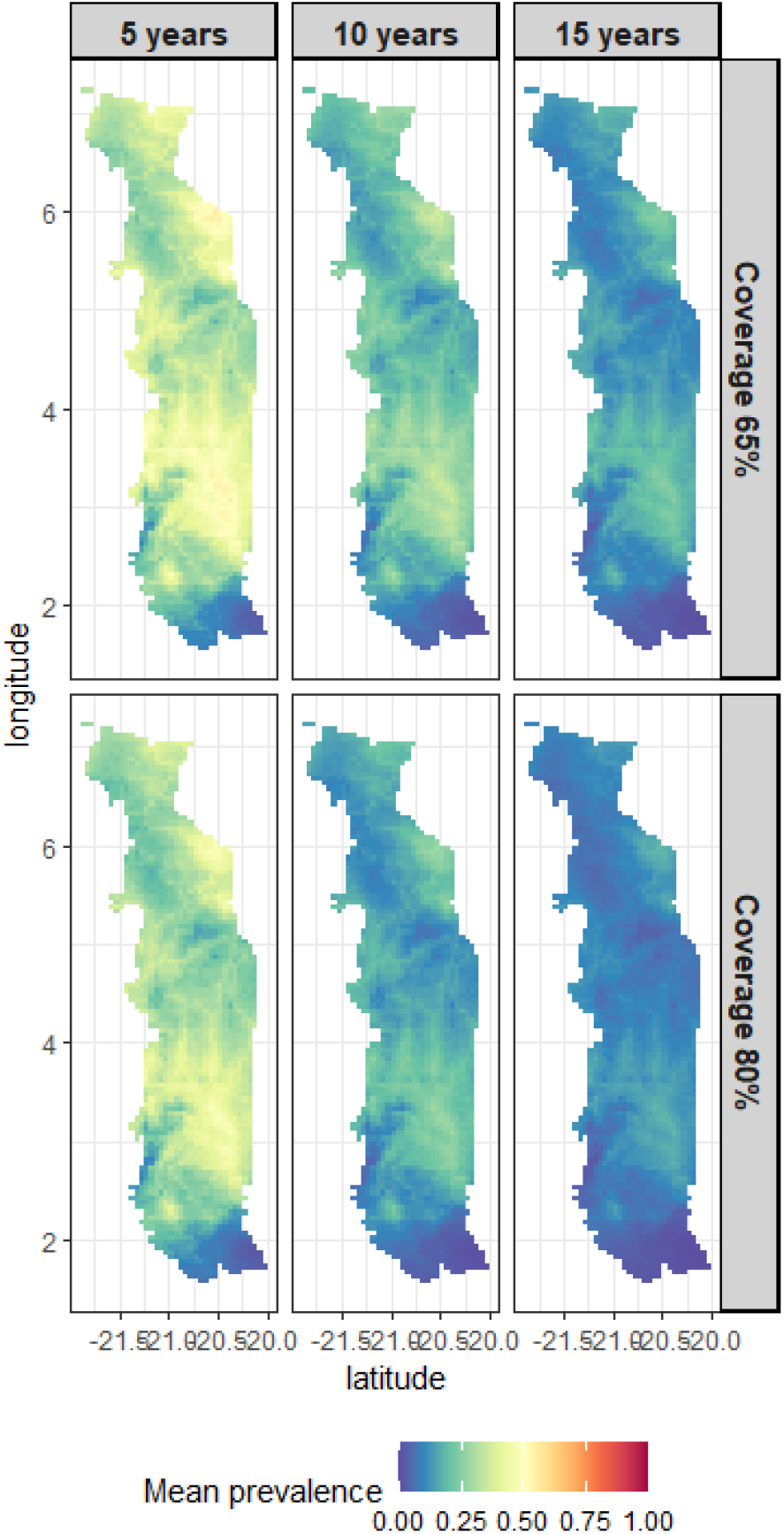
Comparison of the impact of ivermectin coverage levels on elimination of onchocerciasis. The interventions compared are mass drug administration of ivermectin at 65% coverage, and mass drug administration at 80% coverage. Projections run using the EPI-ONCHO model (41).

**Fig 9.**
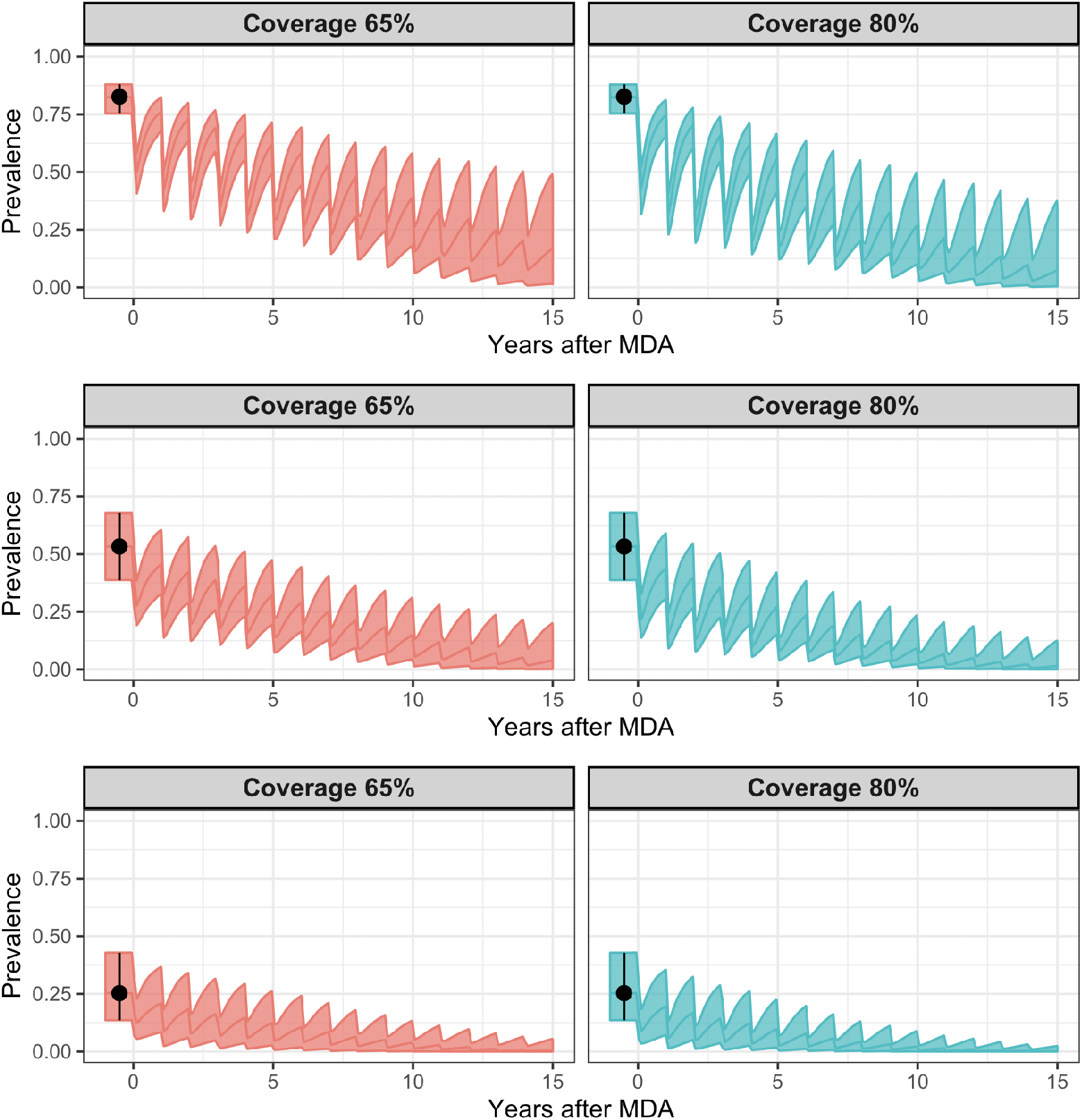
Comparison of two strategies (minimum recommended 65% coverage of total population, and enhanced 80% coverage as an alternative treatment strategy) for onchocerciasis control and elimination at three levels of baseline endemicity (hyper-, meso and hypoendemicity). Projections were simulated using the EPIONCHO model. Projections were simulated using the EPIONCHO model (41). Mapped modelled prevalences are shown in black (4). Figures show mean and 95% inter-quantile range.

### 3.3. HIV infection in Botswana

HIV/AIDS is a sexually transmitted infection with an estimated 6000 new infections occurring globally each day (44). It is a leading cause of disease burden in sub-Saharan Africa (7).

#### 3.3.1. Model and data

We have used the model developed by the Joint United Nations Programme on HIV/AIDS (UNAIDS) called Estimation and Projection Package (EPP) (45). This is a deterministic Susceptible-Infected model with the population divided into three groups: a not-at-risk group, an at-risk group, and an infected group. Prevalence dynamics are defined by four parameters: the start time of the epidemic, *t*_0_; the fraction of the population in the at-risk category at the start of the epidemic, *f*_0_; the rate of infection, *r* (governs the rate at which people in the susceptible population become infected), and the behavioral response, *φ* (this parameter affects the distribution of new entrants to the not at-risk and at-risk categories). We set the initial size of the population to 10,000. For full description of the model see (46).

We have used maps of estimated prevalence of HIV among adults (aged 15–49 years) in sub-Saharan Africa from 2000 to 2017 (7). These maps were at 5km×5km spatial resolution. We have extracted maps for Botswana, which had *I* = 29, 174 pixels per map. However, only maps for mean, lower bound (2.5th percentile) and upper bound (97.5th percentile) were available, which were calculated from the 1,000 posterior draws (7). As this case study is an illustrative example to test how the AMIS framework performs when the dimension of the parameter space gets bigger, we have used these values to re-populate 100 individual maps by assuming that mapped prevalences were drawn from two half-normal distributions with means equal to the mapped mean, and standard deviations as a function of upper/lower bound.

#### 3.3.2. Implementation of AMIS

We set a uniform prior for the rate of infection: *r* ∼ *U* [0, 15], a uniform prior for the fraction of the population in the at-risk category at the start of the epidemic *f*_0_ ∼ *U* [0, 1], a uniform prior for the start time of the epidemic *t*_0_ ∼ *U* [1970, 1990], and a uniform prior for the behavioral response: *φ* ∼ *U* [*−*200, 400]. Here priors for *r, f*_0_ and *t*_0_ are similar to those used in (46), excepting the prior for *φ*, which was set as a normal distribution with mean 0.

We have chosen to use maps of estimated prevalence of HIV in 2000, 2005, 2010 and 2015 (7). Therefor we have *K* = 4 time points to match for each simulated epidemic curve.

#### 3.3.3. Results

The mean mapped prevalence distribution of HIV among adults in Botswana was more homogeneous in comparison to that of ascariasis (Section 3.1) or onchocerciasis (Section 3.2)) (which is not surprising given the exacting ecological requirements for these two helminth infections). HIV prevalence ranged between 0.151 and 0.401 in 2010 (Fig.10(a) (7)). Similar ranges were observed for other years as well: 0.155-0.371 in 2005; 0.151-0.332 in 2010, and 0.137-0.318 in 2015.

Figures 10(b)-(d) show the results of the AMIS framework. The AMIS framework required 17 iterations, i.e., 17,000 parameter vectors (Fig. 10(b)). The algorithm targeted areas of parameter space corresponding to prevalences between 0.1 and 0.4, which is the range of mapped prevalences in 2000. The figure 10 shows a scatter plot and marginal density distribution for two of four parameters: the rate of infection, and the fraction of the population in the at-risk category at the start of the epidemic. Scatter plots for all parameters are shown in supplementary Figure S2. It can be seen that the relationship between parameters and infection prevalence is more complex than in the previous two case.

**Fig 10.**
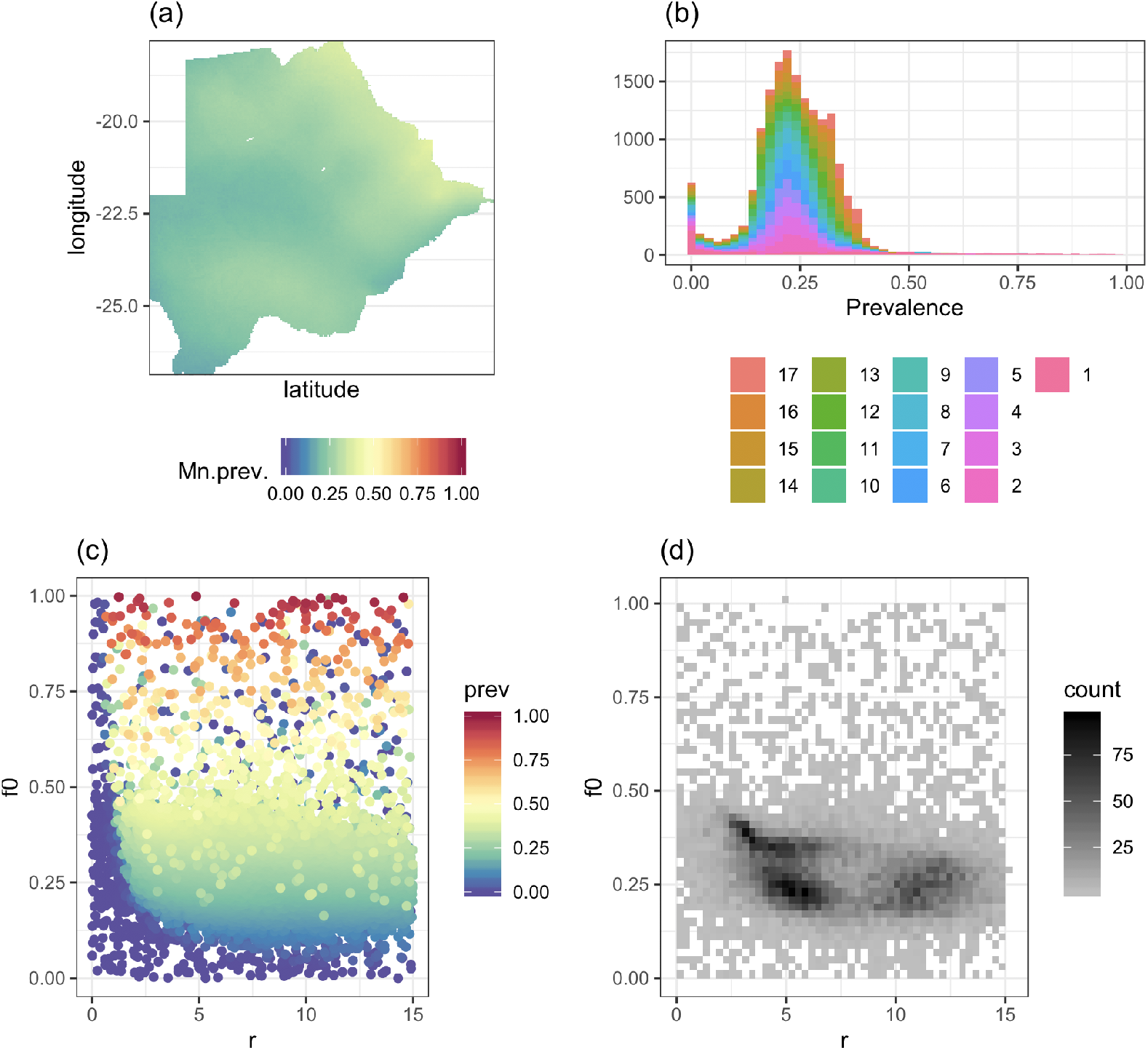
Results for HIV in Botswana. (a) Map of mean prevalences in 2010 (7). The resolution is 5km×5km and M = 100. (b) Histogram of sampled prevalences with colour coding according to the various iterations of the AMIS algorithm. (c) Scatter plot of sampled model parameters and corresponding prevalence. (d) Density of sampled parameters. In (c) and (d), prevalence and density distributions are shown as a function of the rate of infection, r, and the fraction of the population in the at-risk category at the start of the epidemic, f_0_.

We have found high uncertainty about prevalence dynamics before the time of mapped data, i.e. year 2000 (Figure 11). The authors in (46) have restricted prevalence in 1980 to be smaller than 10%. This restriction would be straightforward to implement in our algorithm by introducing an additional map in 1980 with values of prevalence *q*_*i,m*_ ∼ *U* [0, 0.1].

**Fig 11.**
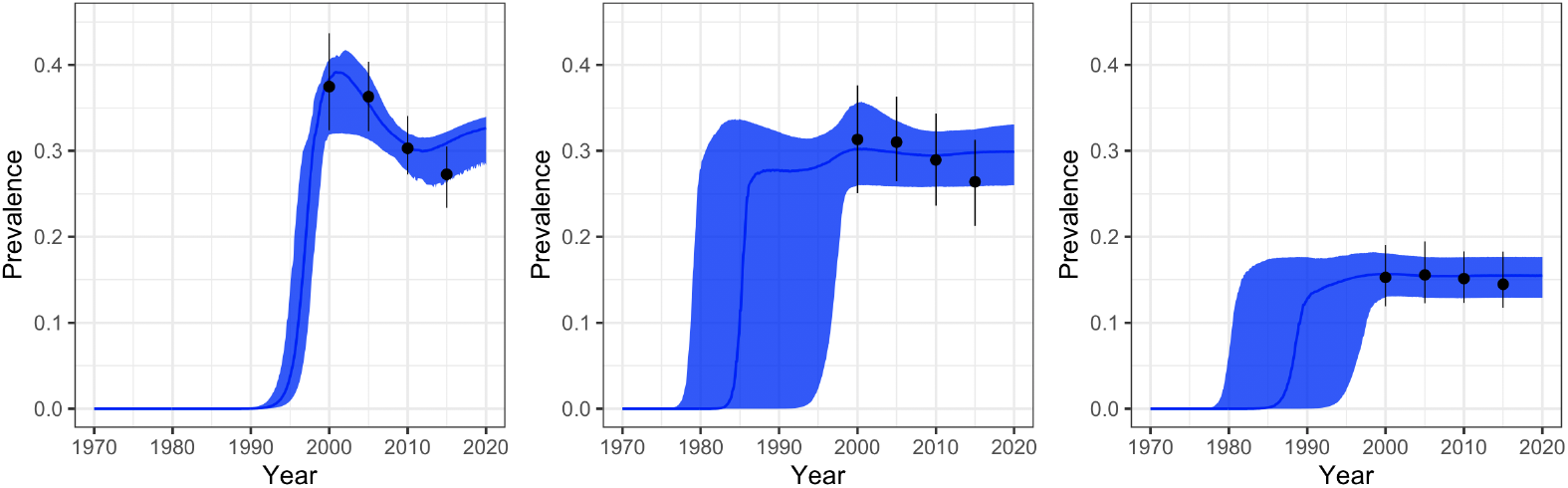
Comparison of mapped and simulated prevalences at three levels of endemicity for HIV in Botswana. Mapped modelled prevalences are shown in black (7). Figures show mean and 95% inter-quantile range.

### 3.4. Malaria in the Democratic Republic of the Congo

Malaria is a serious vector-born disease caused by the parasitic protozoan *Plasmodium* and transmitted among humans via the bites of female *Anopheles* mosquitoes (47). Although malaria infection prevalences have decreased by half in the past decade due to global efforts, there are still an estimated half a million of deaths a year (48). Mathematical modelling of malaria transmission can help to identify localised areas where control effectiveness is failing and provide a rational for planning intervention programmes (49).

#### 3.4.1. Model and data

We have used the OpenMalaria platform to simulate Plasmodium falciparum parasite prevalence (PfPR) under a microscopy-based parasite detection (50). The OpenMalaria platform combines an individual-based stochastic simulation model of malaria in humans with a deterministic model of malaria in mosquitoes. All parameters, except the annual Entomological Inoculation Rate (the number of infectious mosquito bites per person per year), *aEIR*, and the variance between individuals in parasite densities (of asexual stages in erythrocytes), 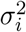, were set as in (51).

Data from PfPR surveys in Africa in 2000 were used to construct a map of PfPR prevalence for children aged 2-10 years at 5km×5km spatial resolution in sub-Saharan Africa (1). We have extracted maps for the DRC, which had *I* = 108, 662 pixels for each map. The data has 100 sample draws of prevalences for each pixel. This case study was different from the onchocerciasis case as we have a large number of pixels, but a small number of maps for each individual pixel.

#### 3.4.2. Implementation of AMIS

We set a uniform prior for the log of annual Entomological Inoculation Rate: log(*aEIR*) ∼ *U* [log(1), log(500)], and a uniform prior for variance in parasite density among individuals, *σ*_*i*,2_ ∼ *U* [0.1, 10]. We set population size to 10,000 and run the model for 100 years to get to an epidemic equilibrium. For details on model simulation see (50). We chose to use maps of the estimated prevalence of malaria in 2005 and 2010. These years were chosen arbitrary, as we were interested to use *K* = 2 time points to match for each simulated epidemic curve. Furthermore, we simulated projections of prevalence up to year 2015 to be able to compare the predicted and mapped prevalences.

In order to simulate malaria transmission from 2000 to 2015, we had to account for the implementation of interventions during these years. Insecticide-treated bednet (ITN) coverage in the DCR in 2015 was downloaded from the *malariaAtlas* Project (MAP) (52). We ignored the coverage of ACTs as this was considered to be low in the DRC (*<* 2% in 2000-2010 and *<* 16% in 2015 (1)). We assumed spatial homogeneity in ITN coverage, which was set to a mean 76%. To calculated annual coverage of ITNs, we used data on total ITNs distributed in the DRC in 2005-2014 (53), and assumed that the average mapped coverage of 76% was achieved by year 2015 (for details see Supplementary section B).

#### 3.4.3. Results

The corresponding mean prevalence map and the results of AMIS parameter estimation are shown in Figure 12. It can be seen from fig. 12(a) that the mean prevalence of malaria in children had a large range of values, i.e. from 0.2 to 0.9. Furthermore, pixels with high/low mean prevalence are spatially clustered. Therefore, it would be expected that many pixels should have very similar prevalence distribution profiles, which would be beneficial for implementing the AMIS algorithm across all pixels of the map.

**Fig 12.**
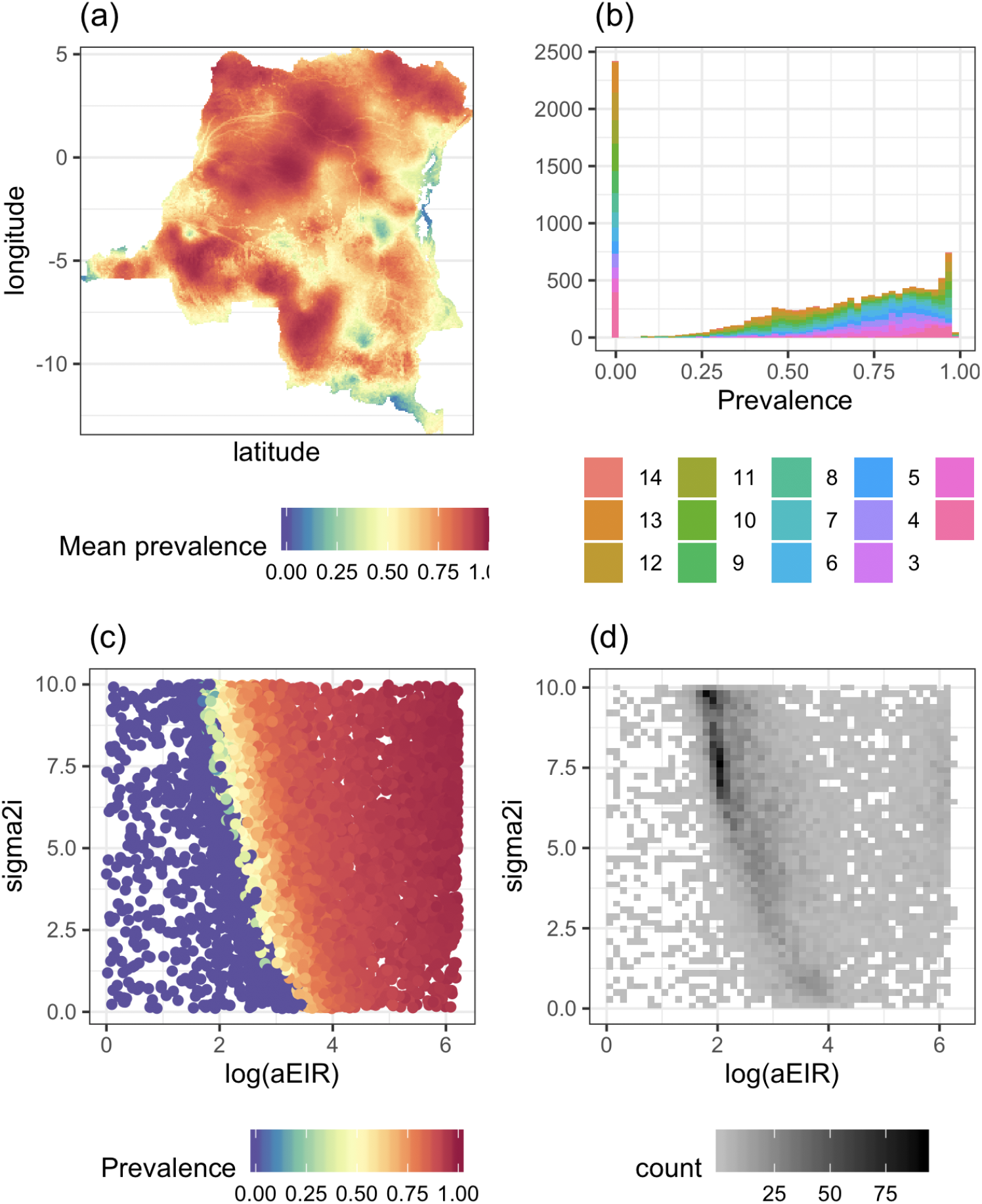
Results for malaria in the Democratic Republic of the Congo. (a) Map of mean prevalences in 2005 (1). The resolution is 5km×5km and M = 100. (b) Histogram of sampled prevalences with colour coding according to the various iterations of the AMIS algorithm. (c) Scatter plot of sampled model parameters and corresponding prevalence. Density of sampled parameters. In (c) and (d), prevalence and density distributions are shown as a function of log annual Entomological Inoculation Rate, aEIR, and the variance between individuals in parasite densities, 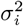. Projections were simulated using the OpenMalaria model (51).

We found that prevalence exhibits a nonlinear relationship with respect to *aEIR* and variabilityvariance in parasite densities among individuals, 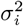, exhibiting a similar tendency to that in ascariasis (Section 3.1) and onchocerciasis case (Section 3.2) (fig. 12(c)). The AMIS framework required 14 iterations to achieve *ESS ≥ ESS*^*R*^ (fig. 12(d)). As can be see from the plot of densities of sampled parameters, AMIS targeted the region of the parameter space corresponding to the largest variation in prevalence (fig. 12(d)).

Figure 13 shows a comparison of mapped and simulated prevalences at three levels of malaria endemicity. Our results demonstrate a sunstantial decline in PfPR prevalence for children aged 2-10 years in 2015, which is over-estimated in comparison to the mapped prevalences. On average, the simulated epidemic curves tend to be lower than mapped prevalences in 2005 as well, which is in contrast to the onchocerciasis case, where simulated baseline prevalences were similar to the mapped prevalences (Fig. 9).

**Fig 13.**
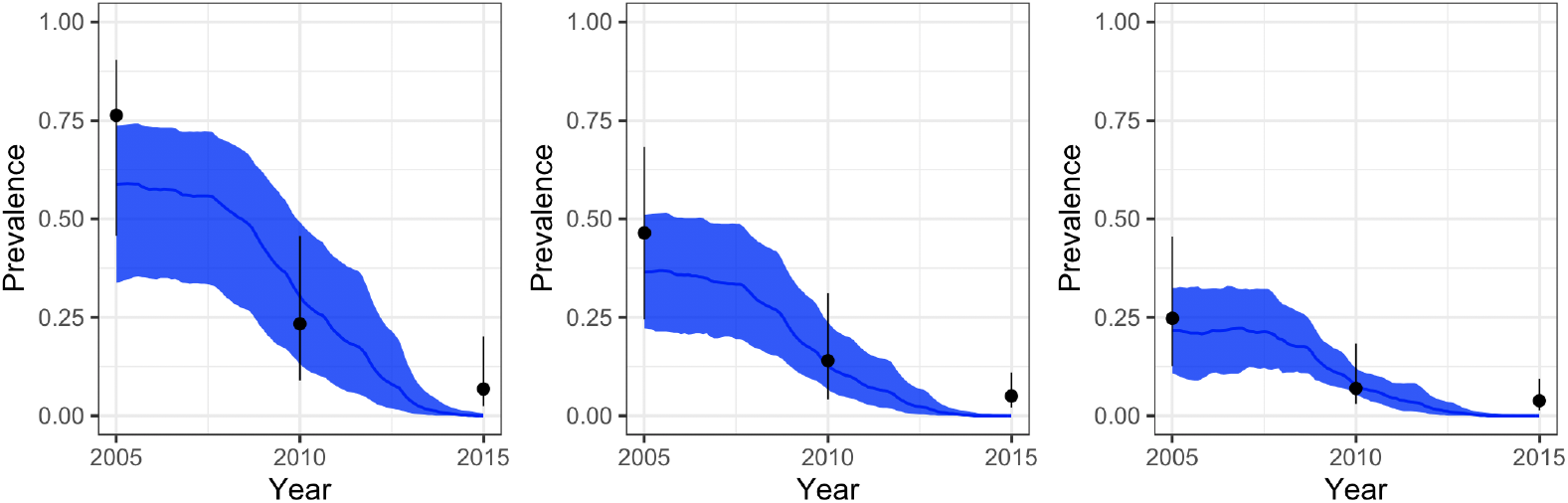
Comparison of mapped and simulated prevalences at three levels of endemicity for Plasmodium falciparum malaria in the DRC. Mapped prevalences are shown in black (1). Projections were simulated using the OpenMalaria model (50). Figures show mean and 95% inter-quantile range.

## 4. Discussion

Our proposed framework allows the distribution of prevalences from high-resolution maps to be projected forward into the future under the transmission dynamics of complex disease transmission models. Below we discuss the benefits and limitations of the methodology.

### 4.1. Parameter estimation based on prevalences at a national level

The diversity of diseases studied and the range of mathematical models used, allowed us to explore the performance of our framework under different conditions. As the results in the previous section showed, the AMIS framework performed well for all four case studies. The proposed algorithm, utilizing the combination of all the samples from multiple proposals, was suitable for the task of parameter estimation based on geospatial maps, as it allowed us to explore complex dependencies between parameters and modelled prevalences with relatively small numbers of transmission model simulations. An alternative strategy where parameters are sampled for each individual pixel independently would require much higher numbers of simulations, as the number of pixels in a map can be large. We used the same simulations for each pixel, with extra computational time required to calculate weights and ESS at each iteration.

### 4.2. Using weights of active pixels

The algorithm we propose has an additional step of calculating the average weight of parameter vectors based on individual weights of active pixels. In section 3.1 we compared this approach with two alternative methods, namely: based on sampling from the prior only, and based on AMIS with |*A*| = *M* for all iterations. As can be seen from Table 1, the method based on weights of active pixels required 2-3 times fewer iterations then the algorithm based on sampling from the prior only or based on AMIS without removing pixels from the active set. The latter algorithm targets simulations towards the pixels that have already been well served by earlier iterations of the algorithm, and not towards the required posterior prevalences for all of the pixels, for example simulated prevalences between 0.5 and 0.75 (Fig. 5).

### 4.3. Influence of size of geospatial maps

In Section 3.1 we found that the number of pixels (i.e. *I* = 11, 369 pixels and *I* = 37, 695 pixels for the prevalence of *Ascaris* in school-aged children in Ethiopia) had little influence on AMIS performance in terms of the number of iterations required (Table 1). The number of individual prevalence maps used for parameter sampling also had a minor influence, with the number of AMIS iterations equal to 8 or 9 when the number of maps varied from 100 to 2000. A slightly lower increase in minimal ESS was observed for *M* = 100, but this did not translate into any additional iterations. This suggests that having a higher number of maps will not require more transmission model simulations, but in fact can decrease this number in comparison to a smaller number of maps. Having a smaller number of maps for each pixel can make it harder to match the sampled prevalences to the distribution of observed prevalences which translates into extra iterations. This may be because there is a spatial correlation between neighbouring pixels, so simulated prevalences close to some observed prevalences at particular pixels will be similar to those in neighbour pixels.

### 4.4. Setting priors of parameters

The AMIS framework applied to transmission models involves defining a prior distribution for model parameters. In most case studies, we used a uniform distribution with parameter ranges informed by the available literature on biological processes governing infection transmission or previous analysis of mathematical models. Our framework allows incorporating specific knowledge of the parameter space, for example excluding particular areas/combinations of parameters that have been deemed biologically unfeasible, or introducing dependencies between parameters and can be informed by additional data. Practical application of this has been demonstrated for *A.lumbricoides* in section 3.1.4, where parameters for the degree of parasite aggregation were sampled from the Gaussian distribution conditioned on the values of mean number of warms.

### 4.5. Impact of intervention programs

In section 3.2 we have sampled parameters for onchocerciasis in Togo, simulated the transmission model further forward in time under two levels of coverage and applied the weights to obtain the spatial distribution of the projections. We found a good agreement between the mapped and simulated baseline prevalences. However, the uncertainty was propagated with time and simulations produce a wide range of future projected prevalences. Uncertainty can be reduced by utilizing post-control maps within the AMIS algorithm.

### 4.6. Comparison to Bayesian melding

A probabilistic approach, called Bayesian melding, combines via logarithmic pooling two priors: one implicit and one explicit, on each input and output (54). Bayesian melding has been proposed and used to account for uncertainty in parameters and model projections for HIV (46). The initial stage of the algorithm required 200,000 combinations of the input parameters from their prior distribution and produced 373 unique epidemic curves fitted to aggregated data on HIV prevalence in urban clinics (46). This data would be equivalent to a single pixel in our case. As shown in Section 3.3, the AMIS algorithm offers improved computation efficiency and higher resolution in comparison to this application of Bayesian melding.

### 4.7. Using post-control maps

For the malaria case study in Section 3.4, our results demonstrate that when sampling parameters using baseline as well as post-control maps, care has to be taken that the simulated epidemiological curves are able to support mapped dynamics. We assumed that 15% coverage of ITNs was achieved by 2010 (Supplementary table S1), but this coverage would not reduce mean prevalence from 0.75 to 0.25, as seen in Figure 13. The AMIS framework produced a trade-off between fitting to maps in 2005 and 2010. However, the schedule of ITNs coverages used within the model was better suitable for pixels with low baseline endemicity. Therefore pixels should be divided into regions with different histories of control. The AMIS framework can then be applied within each region.

### 4.8. Tuning of hyper-parameters

The proposal distribution adapts to the target by locally fitting a mixture component to areas which correspond to pixels with low effective sample sizes. Therefore the performance of the algorithm depends on the choice of threshold *ESS*^*R*^. In our case studies we have set *ESS*^*R*^ = 2000. The cost of AMIS sampling has the following components: fitting a mixture of distributions; sampling parameters from the mixture, performing model simulations for a parameter vector and calculating values of ESS. When the latter two are computationally expensive, lower values of *ESS*^*R*^ would be advisable.

We have used a prior for sampling just for a single iteration. In (16), the authors set half of the iterations to be sampled from the prior. In our case, we don’t know in advance how many iterations will be required to have enough parameter sets to satisfy the condition *min*(*ESS*) *≥ ESS*^*R*^. However, our results for all case studies indicate that having sampled 1000 parameter sets was enough to gauge important regions of parameter space for further analysis, at least in 2 to 4 dimensions. We also had the number of samples per iteration fixed to *N*_*t*_ = 1000. We anticipate that decreasing the value of *N*_*t*_ could lead to a lower number of parameter sets required to achieve min(*ESS*) *≥ ESS*^*R*^, but at a cost of increasing the number of iterations, which will add an additional computational cost due to the requirement to recalculate the weights and ESS after each iteration.

The stability of the algorithm requires that the parameter space has been sufficiently explored in the initial iteration. Low values of ESS would show that there are many parameter samples which carry relatively low weight. However ESS can be deceptively high when all the simulations fall in regions with equally low weight. We avoid this by ensuring that the initial iteration comprises sufficiently many samples so that at least some of them fall in regions with high weight. Depending on mapped prevalences and model behaviour, the distribution of ESS can have a range of values after the first and subsequent iterations. For example in the case of *Ascaris*, we had that min(*ESS*) = 67.4 and max(*ESS*) = 435.1 after the first iteration, which are high in comparison to a number of sampled parameters, i.e. 1000 (Fig. 3).

### 4.9. Alternative AMIS procedures

Recently, a modified AMIS has been proposed, where an importance sampling distribution at iteration *t* is built based on samples from iteration *t −* 1 but weights for all iterations are recalculated after the last iteration (55). A simpler recycling strategy could offer computational savings, but this would lose the advantage of the adaptive nature of the AMIS framework we proposed, i.e. to utilize information on the ESS based on all sampled parameter vectors rather then only the subset from the latest iteration.

A further class of adaptive importance samplers has been proposed, in which the adaptation is driven by independent parallel or interacting Markov chain Monte Carlo chains (56). Parallelisation might be a promising route, for example, by sampling *N*_*t*_*/d* parameter vectors and running the model and projections on *d* parallel units, then combining them into a single iteration and using aggregated parameter vectors to sample next *N*_*t*_*/d* parameter vectors.

## 5. Conclusions

Infectious diseases remain an important health problem worldwide. We have introduced a novel method to sample parameters of disease transmission models given high-resolution prevalence maps. Our strategy of using a mixture of Gaussian distributions based on a transmission model’s similarity to mapped prevalence distributions leads to an efficient exploration of the state space. We have extended the methodology to include maps for multiple time points. In future work, applications of the methodology that account for routine surveillance data would allow greater epidemiological insight into providing tools for policy at a local level, bringing infectious diseases under control, or even setting out the pathway for elimination of transmission.

## Data Availability

https://github.com/rretkute/AMIS_ID

## Acknowledgments

The authors gratefully acknowledge funding of the NTD Modeling Consortium by the Bill and Melinda Gates Foundation [OPP1152057, OPP1053230, OPP1156227]. The views, opinions, assumptions or any other information set out in this article should not be attributed to the Bill and Melinda Gates Foundation or any person connected with the Bill and Melinda Gates Foundation.

## SUPPLEMENTARY MATERIAL

**Supplement: Integrating geostatistical maps and transmission models using adaptive multiple importance sampling** (doi: [COMPLETED BY THE TYPESETTER](https://dx.doi.org/COMPLETED%20BY%20THE%20TYPESETTER); .pdf). We provide additional supporting plots for *Ascaris* and HIV.

The code is available at https://github.com/rretkute/AMISID.

